# Do non-steroidal anti-inflammatory medications or opioids increase the risk of nonunion after surgical and nonsurgical treatment of tibial shaft fractures?

**DOI:** 10.64898/2026.10.29.26364878

**Authors:** Erika Roddy, Laura S. Gold, Patrick J. Heagerty, Janna L. Friedly, Sean D. Rundell, Jeffrey G. Jarvik, Pradeep Suri

## Abstract

**Background:** Nonsteroidal anti-inflammatory drug (NSAID) medications have been implicated in fracture nonunion. However, the evidence to date for this association is limited, due in part to the rarity of the outcome. Prior investigations may not sufficiently account for confounding, limiting the ability to draw accurate conclusions regarding the independent association of NSAID exposure with subsequent nonunion rates. Opioids are often used instead of NSAIDs to treat postoperative pain, but the association between opioid use and nonunion risk is unclear. The purpose of this study was to investigate the impact of NSAID use and opioid use on rates of nonunion in in skeletally mature patients with tibial shaft fractures.

**Patients and Methods:** This was a cohort study using the Merative™ Marketscan® Research databases from January 1 2007 to December 31, 2022, which contain inpatient, outpatient, and pharmacy claims data for roughly 273 million individuals covered by employer-sponsored commercial health insurance throughout all US states and territories. Patients were included if they were greater than 16 and less than 65 years of age with an ICD-9 or ICD-10 diagnosis of a fracture of the tibia or tibia and fibula between 2008 and 2022. 297,334 patients met this inclusion criteria. The exposure for the primary aim was defined as having ≥1 fill of an NSAID in the 0-30 days following fracture diagnosis, as compared to no fills of an NSAID in the 0-30 days following fracture diagnosis. The exposure for the secondary aims was defined as having ≥1 fill of an opioid in the 0-30 days following fracture diagnosis, as compared to no fills of an opioid (aim 2) or NSAID (aim 3) in the 0-30 days following fracture diagnosis. The outcome for all analyses was fracture nonunion, defined as a surgical procedure performed to treat tibial nonunion between 90 and 365 days after fracture diagnosis. Propensity score analysis, including injury, treatment, and demographic variables associated with nonunion, was used to compare nonunion rates between the groups. Cox proportional hazard regression was performed to estimate hazard ratios (HRs) and 95% confidence intervals (95% CIs), adjusted for the variables included in the propensity match.

**Results:** For patients filling NSAIDs in the 30 days following fracture diagnosis, the adjusted Cox proportional hazard ratio (HR) for nonunion was 1.60 (95% CI 1.20-2.15, p=0.002), indicating a greater risk of nonunion for patients who filled a prescription for NSAIDs within 30 days of the fracture compared to those who did not. Similarly, we observed more treatments for nonunion in the group that filled opioids (1.3% versus 0.9%) and a statistically significant greater rate of nonunion for patients filling opioids compared to those not filling opioids (HR 1.41, 95% CI 1.19-1.66, p<0.001). There was no significant difference in the risk of nonunion for patients who filled NSAID prescriptions compared to those who filled opioid prescriptions (HR 0.89, 95% CI 0.45-1.76, p=0.73)

**Conclusion:** In this propensity matched claims analysis of patients aged 17-64 with tibial shaft fractures, filling NSAIDs and filling opioids postoperatively were separately associated with increased risk of surgical treatment for nonunion compared to patients who did not fill NSAIDs or opioids. Patients who filled NSAIDs only had no difference in the rate of surgical treatment for nonunion compared to patients who filled opioids only. Thus, both NSAIDs and opioids are associated with increased risk of nonunion, but neither appears to be of substantially higher risk than the other. Physicians should consider the side effect profiles of both drugs when deciding the type, quantity, and duration of pain medications to prescribe postoperatively.

**Level of Evidence:** level III

## Introduction

Fracture healing is a complex process. When the normal fracture healing response is disrupted or impaired, fracture nonunion can occur.^1^ Nonunion is a devasting complication. Multiple studies have found that the impact of lower extremity fracture nonunion on patients’ physical and mental health is worse than the impact of congestive heart failure, myocardial infarction, end stage hip arthrosis, and a number of other debilitating chronic medical conditions.^2, 3^ Not only does nonunion adversely impact patients’ physical function and mental well-being as well as their ability to return to work, but it also results in significant costs to the health care system.^4, 5^

Thus, efforts to reduce the risk of nonunion are imperative. Non-steroidal anti-inflammatory drug (NSAID) medications are one potential culprit which have been implicated in elevated risk of fracture nonunion. There have been several animal studies demonstrating decreased fracture callus quantity and quality, increased time to union, and increased rates of nonunion following exposure to NSAIDs.^6–8^ However, the evidence in humans is mixed. Some retrospective clinical studies have found an association between NSAID use and increased risk of nonunion, particularly if taken for prolonged periods^9–12^. However, many of these studies control for a very limited range of potential confounding factors, such that the NSAID and non-NSAID groups being compared in these studies may be fundamentally different. Additionally, most prior studies do not use active control groups, which can also predispose to basic differences between intervention and control groups. Many of these studies also examined the use of indomethacin only.^10, 13^ Other clinical studies have shown no adverse effect of NSAIDs on union rate.^9, 14–18^ Overall, prior work may not sufficiently account for confounding, limiting our ability to draw accurate conclusions regarding the independent association of NSAID exposure with subsequent nonunion rates.

Out of concern regarding potential nonunion risk, many if not most orthopaedic surgeons do not prescribe NSAIDs to manage postoperative pain for fracture patients. This may result in overreliance on opioids and contribute to the burden of opioid dependence that exists nationwide. Indeed, although orthopaedic surgeons represent only 2% percent of physicians^19^, they are the third most frequent prescriber of opioids overall and the most frequent amongst surgical subspecialties.^20,21^ NSAIDs have a generally favorable side effect profile compared to opioids.^22^ In non-trauma orthopaedic surgery populations, scheduled NSAID use postoperatively has resulted in improved pain scores, decreased opioid use, and decreased hospital length of study.^22, 23^ Further clarification of the effect of NSAIDs on fracture union is warranted.

The primary aim of this study was to investigate the impact of NSAID use, compared to no NSAID use, on surgical treatment of nonunion in patients with tibial shaft fractures. In order to draw comparisons with opioids, a common alternative to NSAIDs for managing pain after fracture, secondary aims also investigated (a) the impact of opioid use compared to no opioid use on nonunion rates, as well as (b) the impact of NSAID use compared to opioid use.

## Methods

This was a cohort study using the Merative™ Marketscan® Research databases from January 1 2007 to December 31, 2022, which contain inpatient, outpatient, and pharmacy claims data for roughly 273 million individuals covered by employer-sponsored commercial health insurance throughout all US states and territories.^24^ These datasets include International Classification of Diseases (ICD)-9/10 diagnostic codes, Current Procedural Terminology (CPT) codes, filled prescriptions, hospital-administered medications, and demographic information on subscribers. Since the datasets include only de-identified data, it was deemed exempt from requiring Institutional Review Board (IRB) approval by the University of Washington IRB.

### Inclusion criteria

Patients were included if they were greater than 16 and less than 65 years of age with an ICD-9 or ICD-10 diagnosis of a fracture of the tibia or tibia and fibula (see Supplemental Table 1 for codes) between 2008 and 2022. We did not include people age≤16 years because they are not skeletally mature.

### Exclusion criteria

Exclusion criteria included patients with less than 1 year baseline enrollment in the database prior to fracture diagnosis (because we needed to identify diagnoses and prescription fills prior to the fracture), as well as patients with less than 90 days of follow-up (because nonunions are unlikely to be diagnosed within 90 days of injury). Patients with a diagnosis of bone cancer or metastatic cancer (Supplemental Table 1) were similarly excluded because they would be at increased risk of non-union due to their cancers. Patients were excluded if they filled a prescription for NSAIDs prior to fracture diagnosis which had a supply duration indicating the medications could be taken past the fracture diagnosis date.

### Exposures

All exposure variables were defined using data elements prior to and including the 30 days following fracture diagnosis. The exposure for the primary aim was defined as having ≥1 fill of an NSAID in the 0-30 days following fracture diagnosis, as compared to no fills of an NSAID in the 0-30 days following fracture diagnosis (Figure 1). A secondary analysis compared people who filled ≥2 NSAIDs in the 0-30 days following fracture diagnosis with those who did not fill any NSAIDs during this period.

The exposure for the secondary aim comparing opioid use and no opioid use was having ≥1 fill of an opioid, versus no opioid fills, in the 0-30 days after fracture diagnosis. The exposure for the secondary aim comparing NSAID use with opioid use was ≥1 fill of an NSAID only (without filling an opioid prescription), versus ≥1 fill of an opioid only (without filling an opioid prescription), in the 0-30 days after fracture diagnosis.

### Outcomes

The outcome for all analyses was fracture nonunion, defined as a surgical procedure performed to treat tibial nonunion (Supplemental Table 1) after 90 days but within 365 days (to increase the likelihood that the nonunion treatment was treating the index fracture rather than a subsequent one) following initial fracture diagnosis. This was chosen as the study outcome, rather than diagnosis of nonunion alone, as patients who had a ICD-9 or ICD-10 diagnosis of nonunion but did not undergo any procedures to treat the nonunion may represent delayed union rather than true nonunion.

### Covariates

All covariates were defined using data prior to and including the 30 days following fracture diagnosis.

### Contraindications to NSAID use

Comorbidities that may have impacted the likelihood of prescription of NSAIDs were include in the propensity score, including chronic kidney disease, end stage renal disease, acute kidney injury (diagnosis made in the 7 days before/after the fracture diagnosis), diabetes, liver cirrhosis, history of peptic ulcers, gastrointestinal bleeding, congestive heart failure, and active anticoagulation (Supplemental Table 1).

### Comorbidities contributing to increased risk of nonunion

Similarly, comorbidities that have been implicated in increased risk of nonunion were included, including chronic steroid use, obesity, alcohol use disorder, and prior history of fracture nonunion (Supplemental Table 1).

### Fracture/injury/treatment variables

Finally, injury and treatment variables that could affect nonunion rates were included in the analysis, including how the fracture was treated (ICD codes (Supplemental Table 1) for internal fixation, external fixation, casting/nonoperative treatment within 4 weeks of the fracture) and mechanism of injury (ICD codes (Supplemental Table 1) for high energy, low energy, or other/unspecified within 2 weeks of the fracture).

### Other variables

Other variables included in the propensity score analysis included the following sociodemographic and clinical variables: age, sex, Charlson co-morbidity score, geographic region, calendar year, number of pain locations as defined by Mayhew and colleagues, ^25^ opioid use disorder, and other comorbidities including anxiety, depression, chronic pain, coronary artery disease, hypertension, cerebrovascular disease, inflammatory arthritis, peripheral vascular disease (Supplemental Table 1). Finally, we considered pain medication fills in the year prior to the fracture in order to further account for treatments for pain as proxies of baseline pain severity. This aspect is critical to account for the possibility that those in the non-NSAID comparator group may have had lower pain severity than those in the NSAID group, which may be precisely why they had not been prescribed NSAIDs. To this end we accounted for the covariates of having an opioid fill (yes/no), the number of opioid fills, opioid morphine milligram equivalents (MMEs), duration of opioid fills (sum of the days’ supplies), an NSAID fill (yes/no), number of NSAID fills, a muscle relaxant fill (yes/no), an acetaminophen fill (yes/no), a gabapentin fill (yes/no), and a duloxetine fill (yes/no) in the year prior to fracture. We also included yes/no indicators of whether people had pain medication fills in the 0-30 days after the fracture for the following medications: muscle relaxant, acetaminophen, gabapentin, and duloxetine.

### Statistical analysis

In addition to the propensity score, matching was used to minimize confounding by factors selected *a priori* based on their potentially greater conceptual importance as predictors of nonunion risk. Patients were matched on tobacco use in the year prior to fracture; having an open fracture; presence of open reduction and internal fixation surgery [ORIF] in days 0-7 after fracture diagnosis (vs no ORIF in days 0-7) and days 8-30 (vs no ORIF in days 8-30); and occurrence of a surgical site infection from days 0-30. Tobacco use, open fracture, and surgical site infection were chosen as matching variables as they are consistently associated with the development of tibial nonunion.^26–32^ Timing of surgery was a matching variable in order to accurately assess duration of pain medication use after surgery. In order to compare patients receiving similar amounts of pain medication, patients were matched on quartiles of morphine milligram equivalents (MMEs) prescribed from 0-30 days, as well as on number opioid fills from 0-30 days, categorized as 0 fills, 1 fill, or >1 fill.

After matching, descriptive statistics were calculated (numbers, proportions, medians, interquartile ranges (IQRs) and standardized mean differences to ensure adequacy of matching. Kaplan Meier curves were plotted to evaluate the proportional hazards assumption, which was met for all analyses. Cox proportional hazard regression was then performed to estimate hazard ratios (HRs) and 95% confidence intervals (95% CIs), adjusted for the variables included in the propensity match. In the primary analysis, people who filled NSAIDs were compared to those who did not. In a secondary analysis, people who filled more than 1 NSAID prescription within 30 days were compared to those who did not fill any NSAID prescriptions. Statistical significance was set as p<0.05. All analysis was performed using SAS software version 9.4 (Cary, North Carolina).

The same analytic approach was then repeated for the secondary aims comparing (a) people who filled opioids from 0-30 days to those who did not fill opioids, and (b) people who filled NSAIDs only (but did not fill opioids) with those who filled opioids only (but did not fill NSAIDs) The latter analysis comparing people who filled NSAIDs vs. opioids is an example of an active control design in that both treatment groups were defined based on the need for pain medications, which may help to mitigate between-group differences. This comparison may also be more clinically relevant, given that most patients are provided some form of pain control after sustaining a tibial shaft fracture.

A *pre hoc* power analysis was performed. Based on prior administrative claims studies finding nonunion rates of approximately 1% in tibial shaft fractures, and assuming a relevant difference would be a 100% increase in risk of nonunion in those who filled NSAIDs (or 2% absolute risk), 4636 patients would be needed in each group, assuming a beta of 0.2 and alpha of 0.05.

## Results

### NSAID vs. no NSAIDs

A total of 297,334 patients were identified with a diagnosis of a tibial shaft fracture between 2008-2022. After exclusion criteria were applied, a total of 80,417 patients remained (Figure 1). Of these, 71, 778 (11%) filled a prescription for NSAIDs within 30 days of fracture diagnosis. After propensity matching was applied, 8,446 patients were left in each of the groups of people who filled NSAIDs or did not fill NSAIDs within 30 days of fracture.

Table 1 demonstrates the baseline characteristics of the matched and unmatched cohorts. Prior to matching, people who filled NSAIDs within 30 days of fracture differed across many characteristics from those who did not, particularly with regard to pain medication fills in the 30 days after the fracture. After matching, the two groups were quite similar (SMDs <0.10 for all variables)

**Table 1.** Baseline characteristics of the unmatched and matched cohort comparing those who filled NSAIDs within 30 days of tibia fracture versus those who did not.

| **Variable (# (%)) unless otherwise specified)** | **Unmatched Cohort** |  |  | **Matched Cohort** |  |  |
|---|---|---|---|---|---|---|
|  | **No NSAID (n=71,778)** | **NSAID (n=8,639)** | **SMD** | **No NSAID (n=8446)** | **NSAID (n=8446)** | **SMD** |
| **Demographics** |  |  |  |  |  |  |
| Age in years (median (IQR)) | 47 (32, 56) | 44 (31, 54) | -0.09 | 44 (29, 54) | 44 (31, 54) | 0.03 |
| Male | 36,611 (51.0%) | 4,345 (50.3%) | -0.01 | 4,240 (50.2%) | 4,246 (50.3%) | 0.00 |
| Female | 35,167 (49.0%) | 4,294 (49.7%) |  | 4,206 (49.8%) | 4,200 (49.7%) |  |
| Geographic region |  |  |  |  |  |  |
| North Central | 11,698 (16.3%) | 1,221 (14.1%) | -0.06 | 1,219 (14.4%) | 1,209 (14.3%) | 0.00 |
| South | 17,687 (24.6%) | 1,801 (20.8%) | -0.09 | 1,763 (20.9%) | 1,789 (21.2%) | 0.01 |
| West | 28,726 (40.0%) | 4,000 (46.3%) | 0.13 | 3,923 (46.4%) | 3,939 (46.6%) | 0.00 |
| Northeast | 12,928 (18.0%) | 1,526 (17.7%) | -0.01 | 1,541 (18.2%) | 1,509 (17.9%) | -0.01 |
| Year of fracture exposure (median (IQR)) | 2012 (2010, 2015) | 2013 (2010, 2017) | 0.26 | 2013 (2011, 2017) | 2013 (2010, 2017) | -0.02 |
| **Exact Matching Variables** |  |  |  |  |  |  |
| History of tobacco use | 4,082 (5.7%) | 544 (6.3%) | 0.03 | 498 (5.9%) | 498 (5.9%) | 0.00 |
| Open fracture | 10,627 (14.8%) | 892 (10.3%) | -0.14 | 855 (10.1%) | 855 (10.1%) | 0.00 |
| Surgical site infection within 30 days | 377 (0.5%) | 43 (0.5%) | 0.00 | 29 (0.3%) | 29 (0.3%) | 0.00 |
| MME from 0-30 days |  |  |  |  |  |  |
| 0 | 28,690 (40.0%) | 2,022 (23.4%) | -0.36 | 1,987 (23.5%) | 1,987 (23.5%) | 0.00 |
| >0 to <45 | 13,627 (19.0%) | 2,744 (31.8%) | 0.30 | 2,676 (31.7%) | 2,676 (31.7%) | 0.00 |
| ≥45 to <75 | 14,156 (19.7%) | 2,083 (24.1%) | 0.11 | 2,031 (24.0%) | 2,031 (24.0%) | 0.00 |
| ≥75 | 15,305 (21.3%) | 1,790 (20.7%) | -0.01 | 1,752 (20.7%) | 1,752 (20.7%) | 0.00 |
| # of opioid fills from 0-30 days |  |  |  |  |  |  |
| 0 | 30,197 (42.1%) | 2,070 (24.0%) | -0.39 | 2,032 (24.1%) | 2,032 (24.1%) | 0.00 |
| 1 | 16,748 (23.3%) | 2,616 (30.3%) | 0.16 | 2,558 (30.3%) | 2,558 (30.3%) | 0.00 |
| >1 | 24,833 (34.6%) | 3,953 (45.8%) | 0.23 | 3,856 (45.7%) | 3,856 (45.7%) | 0.00 |
| ORIF from 0-7 days | 13,671 (19.0%) | 1,391 (16.1%) | -0.08 | 1,349 (16.0%) | 1,349 (16.0%) | 0.00 |
| ORIF from 8-30 days | 1,193 (1.7%) | 171 (2.0%) | 0.02 | 148 (1.8%) | 148 (1.8%) | 0.00 |
| **Diagnoses indicating contraindication to NSAID use within year prior to fracture** |  |  |  |  |  |  |
| Chronic kidney disease | 1,332 (1.9%) | 67 (0.8%) | -0.09 | 61 (0.7%) | 64 (0.8%) | 0.00 |
| End stage renal disease | 398 (0.6%) | 10 (0.1%) | -0.08 | 11 (0.1%) | 8 (0.1%) | -0.01 |
| Acute kidney injury (+/- 7 days of fracture) | 960 (1.3%) | 45 (0.5%) | -0.09 | 35 (0.4%) | 44 (0.5%) | 0.02 |
| Diabetes | 7,775 (10.8%) | 741 (8.6%) | -0.08 | 676 (8.0%) | 719 (8.5%) | 0.02 |
| Cirrhosis | 1,895 (2.6%) | 206 (2.4%) | -0.02 | 194 (2.3%) | 199 (2.4%) | 0.00 |
| Peptic ulcer disease | 386 (0.5%) | 26 (0.3%) | -0.04 | 24 (0.3%) | 25 (0.3%) | 0.00 |
| GI bleed | 494 (0.7%) | 26 (0.3%) | -0.06 | 25 (0.3%) | 26 (0.3%) | 0.00 |
| Congestive heart failure | 1,619 (2.3%) | 109 (1.3%) | -0.08 | 102 (1.2%) | 108 (1.3%) | 0.01 |
| Active anticoagulation | 2,979 (4.2%) | 166 (1.9%) | -0.13 | 126 (1.5%) | 160 (1.9%) | 0.03 |
| **Diagnoses indicating increased risk of nonunion** |  |  |  |  |  |  |
| Chronic steroid use | 1,146 (1.6%) | 113 (1.3%) | -0.02 | 104 (1.2%) | 107 (1.3%) | 0.00 |
| Obesity | 5,130 (7.1%) | 699 (8.1%) | 0.04 | 681 (8.1%) | 671 (7.9%) | 0.00 |
| Alcohol use disorder | 224 (0.3%) | 21 (0.2%) | -0.01 | 26 (0.3%) | 20 (0.2%) | -0.01 |
| History of tibial fracture nonunion | 568 (0.8%) | 29 (0.3%) | -0.06 | 20 (0.2%) | 28 (0.3%) | 0.02 |
| **Fracture/treatment characteristics** |  |  |  |  |  |  |
| Internal fixation | 14,842 (20.7%) | 1,562 (18.1%) | -0.07 | 1,501 (17.8%) | 1,502 (17.8%) | 0.00 |
| External fixation | 1,979 (2.8%) | 165 (1.9%) | -0.06 | 163 (1.9%) | 157 (1.9%) | -0.01 |
| Casting/nonoperative treatment | 2,264 (3.2%) | 300 (3.5%) | 0.02 | 274 (3.2%) | 296 (3.5%) | 0.01 |
| Mechanism of injury: High | 12,162 (16.9%) | 1,560 (18.1%) | 0.03 | 1,487 (17.6%) | 1,514 (17.9%) | 0.01 |
| Mechanism of injury: Low | 5,799 (8.1%) | 889 (10.3%) | 0.08 | 858 (10.2%) | 865 (10.2%) | 0.00 |
| Mechanism of injury: Other | 54,991 (76.6%) | 6,360 (73.6%) | -0.07 | 6,264 (74.2%) | 6,230 (73.8%) | -0.01 |
| **Other diagnoses within year prior to fracture** |  |  |  |  |  |  |
| Charlson co-morbidity score (median (IQR)) | 0 (0, 1) | 0 (0, 0) | -0.13 | 0 (0, 0) | 0 (0, 0) | 0.02 |
| 0 | 52,607 (73.3%) | 6,611 (76.5%) | 0.07 | 6,564 (77.7%) | 6,482 (76.7%) | -0.02 |
| 1-2 | 14,357 (20.0%) | 1,653 (19.1%) | -0.02 | 1,560 (18.5%) | 1,606 (19.0%) | 0.01 |
| 3-4 | 3,086 (4.3%) | 285 (3.3%) | -0.05 | 234 (2.8%) | 270 (3.2%) | 0.03 |
| 5+ | 1,728 (2.4%) | 90 (1.0%) | -0.11 | 88 (1.0%) | 88 (1.0%) | 0.00 |
| Generalized anxiety disorder | 2,116 (2.9%) | 357 (4.1%) | 0.06 | 361 (4.3%) | 345 (4.1%) | -0.01 |
| Mood disorder | 5,492 (7.7%) | 750 (8.7%) | 0.04 | 730 (8.6%) | 721 (8.5%) | 0.00 |
| Chronic pain | 2,105 (2.9%) | 274 (3.2%) | 0.01 | 227 (2.7%) | 255 (3.0%) | 0.02 |
| Coronary artery disease | 2,639 (3.7%) | 219 (2.5%) | -0.07 | 190 (2.2%) | 213 (2.5%) | 0.02 |
| Hypertension | 17,761 (24.7%) | 2,077 (24.0%) | -0.02 | 1,979 (23.4%) | 2,013 (23.8%) | 0.01 |
| Cerebrovascular disease | 1,877 (2.6%) | 156 (1.8%) | -0.06 | 129 (1.5%) | 149 (1.8%) | 0.02 |
| Inflammatory arthritis | 1,219 (1.7%) | 165 (1.9%) | 0.02 | 169 (2.0%) | 157 (1.9%) | -0.01 |
| Peripheral vascular disease | 1,720 (2.4%) | 129 (1.5%) | -0.07 | 101 (1.2%) | 122 (1.4%) | 0.02 |
| # of clusters of pain (median (IQR)) | 2 (0, 3) | 1 (0, 3) | -0.04 | 1 (0, 3) | 1 (0, 3) | 0.01 |
| Opioid use disorder | 4,473 (6.2%) | 512 (5.9%) | -0.01 | 459 (5.4%) | 493 (5.8%) | 0.02 |
| **Pain medication fills in year prior to fracture** |  |  |  |  |  |  |
| Opioid fill | 26,800 (37.3%) | 3,021 (35.0%) | -0.05 | 3,054 (36.2%) | 2,921 (34.6%) | -0.03 |
| # opioid fills (among those with ≥1) (median (IQR)) | 2 (1, 3) | 2 (1, 4) | 0.04 | 2 (1, 3) | 2 (1, 4) | 0.07 |
| Morphine milligram equivalents (among those with ≥1 fill) (median (IQR)) | 40 (29, 62) | 38 (25, 58) | -0.05 | 38 (26, 56) | 38 (25, 58) | 0.03 |
| Duration of opioid (sum of days' supplies among those with ≥1) (median (IQR)) | 8 (4, 23) | 7 (3, 26) | 0.00 | 7 (3, 20) | 7 (3, 25) | 0.06 |
| NSAID fill | 12,683 (17.7%) | 2,572 (29.8%) | 0.29 | 2,269 (26.9%) | 2,468 (29.2%) | 0.05 |
| # of NSAID fills (among those with ≥1) (median (IQR)) | 1 (1, 2) | 2 (1, 3) | 0.38 | 1 (1, 3) | 1 (1, 3) | 0.01 |
| Muscle relaxant fill | 7,633 (10.6%) | 1,097 (12.7%) | 0.06 | 989 (11.7%) | 1,044 (12.4%) | 0.02 |
| Acetaminophen fill | 1,022 (1.4%) | 175 (2.0%) | 0.05 | 166 (2.0%) | 168 (2.0%) | 0.00 |
| Gabapentin fill | 3,309 (4.6%) | 433 (5.0%) | 0.02 | 368 (4.4%) | 406 (4.8%) | 0.02 |
| Duloxetine fill | 1,943 (2.7%) | 224 (2.6%) | -0.01 | 202 (2.4%) | 212 (2.5%) | 0.01 |
| **Pain medication fills in 30 days after fracture** |  |  |  |  |  |  |
| Opioid fill | 41,581 (57.9%) | 6,569 (76.0%) | 0.39 | 6,414 (75.9%) | 6,414 (75.9%) | 0.00 |
| Muscle relaxant fill | 5,532 (7.7%) | 1,138 (13.2%) | 0.18 | 1,083 (12.8%) | 1,087 (12.9%) | 0.00 |
| Acetaminophen fill | 318 (0.4%) | 100 (1.2%) | 0.08 | 82 (1.0%) | 94 (1.1%) | 0.01 |
| Gabapentin fill | 3,046 (4.2%) | 762 (8.8%) | 0.19 | 707 (8.4%) | 718 (8.5%) | 0.00 |
| Duloxetine fill | 837 (1.2%) | 139 (1.6%) | 0.04 | 119 (1.4%) | 126 (1.5%) | 0.01 |

Abbreviations
IQR: Interquartile range
MME: Morphine milligram equivalents
NSAID: Non-steroidal anti-inflammatory drugs
ORIF: Open reduction and internal fixation
SMD: Standardized Mean Difference. Values >0.10 could indicate the matching is insufficient

A total of 194 patients (1%) underwent a procedure to treat nonunion within 1 year of injury, including 72 patients (0.9%) in the no-NSAID group and 122 patients (1.4%) in the NSAID group (Table 2). The adjusted Cox proportional hazard ratio (HR) for nonunion between 31 and was 1.60 (95% CI 1.20-2.15, p=0.002) for patients filling NSAIDs in the 30 days following fracture diagnosis, indicating a greater risk of nonunion for patients who filled a prescription for NSAIDs within 30 days of the fracture compared to those who did not (Supplemental Figure 1 and Figure 2).

**Table 2.** Outcomes and reasons for end of follow-up in the comparison of those who filled NSAIDs within 30 days versus those who did not.

|  | **No NSAID**<br>**n=8446** | **NSAID**<br>**n=8446** |
|---|---|---|
| Procedure to treat non-union (primary outcome) | 72 (0.9%) | 122 (1.4%) |
| Days to procedure to treat non-union (median, IQR) | 201 (146, 273) | 196 (152, 246) |
| **Reason for end of follow-up** |  |  |
| Enrollment ended before 1 year | 2,275 (26.9%) | 2,175 (25.8%) |
| Reached 1 year without outcome | 6,067 (71.8%) | 6,102 (72.2%) |
| Study ended (Dec 31, 2022) | 32 (0.4%) | 47 (0.6%) |
| Non-union treatment | 72 (0.9%) | 122 (1.4%) |

In secondary analyses to compare patients who filled more than 1 prescription for NSAIDs within 30 days of fracture with those who did not fill any NSAIDs within 30 days of fracture, 996 patients remained in each group after matching (Supplemental Figure 8) and the groups were well matched (Supplemental Table 6). We observed a greater proportion of nonunions in the group that filled more than 1 prescription of NSAIDs (1.8% vs 0.8%; Supplemental Table 7) and non-statistically significantly increased risk of nonunion for patients filling more than 1 prescription, with a hazard ratio of 2.30 (95% CI 0.91-5.83), p=0.08 (Supplemental Figures 9 and 10).

### Opioids vs no opioids

After applying exclusion criteria, 76,597 patients remained (Supplemental Figure 2). Of these, 44,188 patients filled a prescription for opioids in the 0-30 days within 30 days of fracture, while 32,409 did not. After matching, 27,046 patients were left in each group and the groups were well-matched (Supplemental Table 2). We observed more treatments for nonunion in the group that filled opioids (1.3% versus 0.9%; Supplemental Table 3) and a statistically significant greater rate of nonunion for patients filling opioids compared to those not filling opioids (HR 1.41, 95% CI 1.19-1.66, p<0.001) (Supplemental Figures 3 and 4).

### NSAIDs vs. opioids

After applying exclusion criteria, 46,823 patients were identified who filled either opioids or NSAIDs, including 2,731 patients prescribed NSAIDs only and 44,092 patients who filled opioids only (Supplemental Figure 5). After applying propensity analysis matching, 2,666 patients were left in each group and the groups were well-matched (Supplemental Table 4). We observed the same proportion (0.7%) of people with nonunion in each group (Supplemental Table 5) and no significant difference in the risk of nonunion for patients who filled NSAID prescriptions compared to those who filled opioid prescriptions (HR 0.89, 95% CI 0.45-1.76, p=0.73) (Supplemental Figures 6 and 7).

## Discussion

In this matched propensity claims analysis of patients aged 17-64 with tibial shaft fractures, filling (vs. not filling) NSAIDs was associated with greater subsequent risk of surgical treatment for nonunion when adjusting for a wide range of potential confounders. Similarly, filling (vs not filling) opioids was associated with a greater risk of surgical treatment for nonunion. Finally, there were no significant differences in rates of surgical treatment for nonunion between patients who filled NSAIDs only and those who filled opioids only.

The mechanism by which NSAIDs may inhibit bone healing has been well-demonstrated by in-vitro studies. Prostaglandin E2 (PGE2) plays a key role in osteoblast regulation and bone formation. NSAIDs inhibit the enzymes cyclo-oxygenase-1 (COX-1) and cyclo-oxygenase-2 (COX-2), which in turn inhibit PGE2. However, although several animal studies have found increased risk of poor callus formation and nonunion with the administration of NSAIDs, the evidence in human studies is decidedly mixed. ^9–12^ ^13–17^ The findings in this study contradict the results from two other recent administrative database type studies on the impact of NSAIDs on fracture healing,^9, 15^ which found that short-term prescription of NSAIDs were not associated with increased risk of fracture nonunion.

This difference may be due to a number of differences between the present study and the aforementioned ones. Firstly, in this study exact matching was performed on a number of patient, injury, and treatment-related factors that are strongly implicated in development of tibial nonunions (open fracture, tobacco use, and surgical site infection), and propensity score analysis was used to account for a large number of other demographic, comorbid and injury conditions that could impact bone healing. Further, patients’ baseline levels of pain and pain medication usage were accounted for, to address the issue that patients with higher pre-existing levels of pain may be more likely to fill pain medications including NSAIDs. Patients with chronic opioid use have also been found to have higher rates of nonunion^12^, and thus matching on baseline pain medication usage also helped isolate the potential impact of acute NSAID exposure. These two design features provide more rigorous adjustment for potential confounding than prior studies using administrative or electronic health record data to examine these research questions. Finally, only tibial shaft fractures were included in order to have a more homogenous population. Fractures in different locations can behave quite differently based on amount of surrounding soft tissues and propensity for open fracture, wound complications, and nonunion. Most prior studies examining the impact of NSAIDs on fracture healing included multiple types of long-bone fractures.^9, 15^ Although these studies included fracture location as a covariate in their analyses, it is possible that residual confounding by fracture location remained.

As others have demonstrated, this study found that filling opioids was associated with increased risk of nonunion.^9, 12^ Although the study design attempted to account for a wide range of potential confounding factors including open fracture, mechanism of injury, and type of treatment (surgical vs nonoperative), it is possible that fills of opioids is a surrogate for more severe injuries with higher risk of nonunion.

The risk of nonunion for people who filled NSAIDs compared to those who did not (HR 1.60) was generally similar to the risk of nonunion for people who filled opioids vs those who did not (HR 1.41). Thus, although NSAIDs may increase the risk of nonunion, they appear to do so at a rate similar to opioids. Further supporting this equivalency between NSAIDs and opiates in terms of nonunion risk are the results of the analysis directly comparing people who filled NSAIDs (only) to those who filled opioids (only). In this analysis, there was no difference in nonunion risk between the two groups (HR 0.89 with NSAIDs as compared to opioids, p=0.73), although this analysis was likely underpowered as its per-group sample size (n=2666) was lower than the minimum sample size suggested by our power analysis (n=4636). Nevertheless, our study findings suggest the possibility that, given the increased adverse event profile associated with opioids as compared to NSAIDs, including but not limited to the potential for opioid tolerance and abuse, ^22, 33–39^ physicians may consider attempting to replace opiates with NSAIDs in appropriate patients, given the similar risk of nonunion.

Although attempts were made to control for injury-related factors that may be associated with increased risk of nonunion, including energy of injury, open fracture, and type of treatment, it is possible that residual confounding remained. Filling prescriptions for NSAIDs or opioids may be a surrogate for people with more severe injuries with more resulting pain, who are at increased baseline risk for nonunion. It may be that residual injury-related characteristics account for the increased risk seen with both NSAIDs (vs. no NSAIDs) and opioids (vs. no opioids), but are not seen when comparing NSAIDs alone vs opioids alone since these groups may have more similar pain characteristics.

The rate of nonunion at 1 year was lower in this cohort compared to some other studies (5%-10%).^11, 40, 41^ However, the rate was similar to that reported in other administrative database claims studies.^9, 15^ These findings may be the result of differences in patient population as well as in the criteria used to ascertain nonunion amongst the various studies. The definition of nonunion used in this study as well as others with similar nonunion rates was more stringent: instead of defining nonunion only by diagnosis codes, we required people to have undergone procedures to treat nonunion as our primary outcome measure. Tibial shaft nonunions are greatly disabling and the majority of patients with true nonunion are likely to go on to receive a procedure for nonunion repair.

Limitations of this study include those inherent to all administrative claims studies. Diagnosis coding can be inaccurate. This study was only able to ascertain whether patients filled NSAIDs and opioids, and not the actual specifics of whether or how these medications were used. This study also did not account for patients who may have taken NSAIDs over the counter. Although patients in both groups are subject to this potential bias, patients who did not fill a NSAID prescription may be more likely to have used over the counter NSAIDs. This represents a potential bias towards the null hypothesis. On the other hand, the outcome variables of surgical treatment of nonunion is likely to be accurate given that the coding of these procedures has major financial implications for medical centers and providers. Finally, the patient population in this study is a privately insured population aged 17-64 years. This group may have some inherent differences compared to uninsured or publicly insured populations, as well as compared to patients 65 years of age or greater, and our findings may not be generalizable to all patients with tibia fractures.

## Conclusions

In conclusion, in this propensity matched claims analysis of patients aged 17-64 with tibial shaft fractures, filling NSAIDs and filling opioids postoperatively were separately associated with increased risk of surgical treatment for nonunion compared to patients who did not fill NSAIDs or opioids. Patients who filled NSAIDs only had no difference in the rate of surgical treatment for nonunion compared to patients who filled opioids only. Thus, both NSAIDs and opioids are associated with increased risk of nonunion, but neither appears to be of substantially higher risk than the other. Physicians should consider the side effect profiles of both drugs when deciding the type, quantity, and duration of pain medications to prescribe postoperatively.

## Data Availability

All data produced in the present study are available upon reasonable request to the authors

---

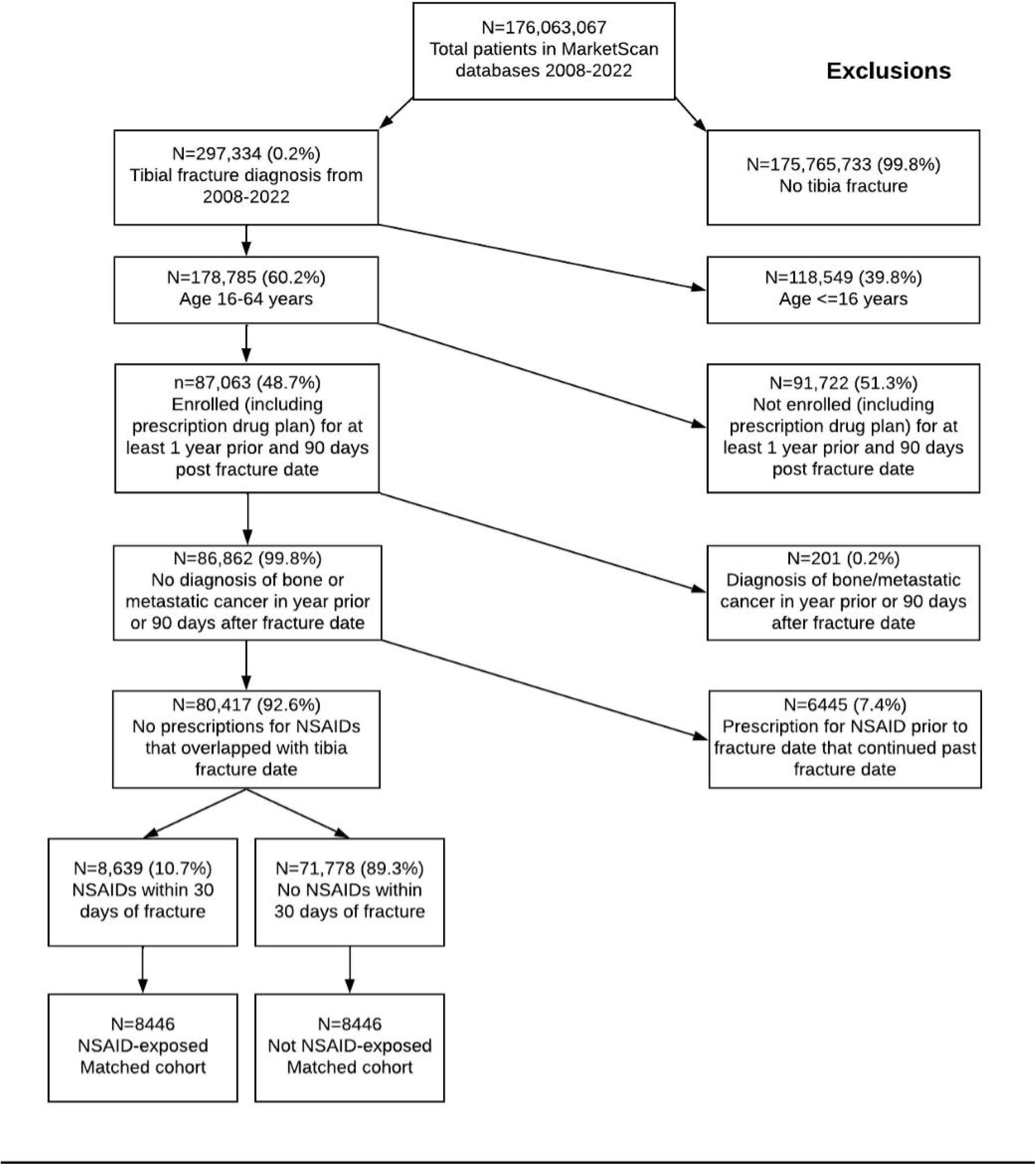

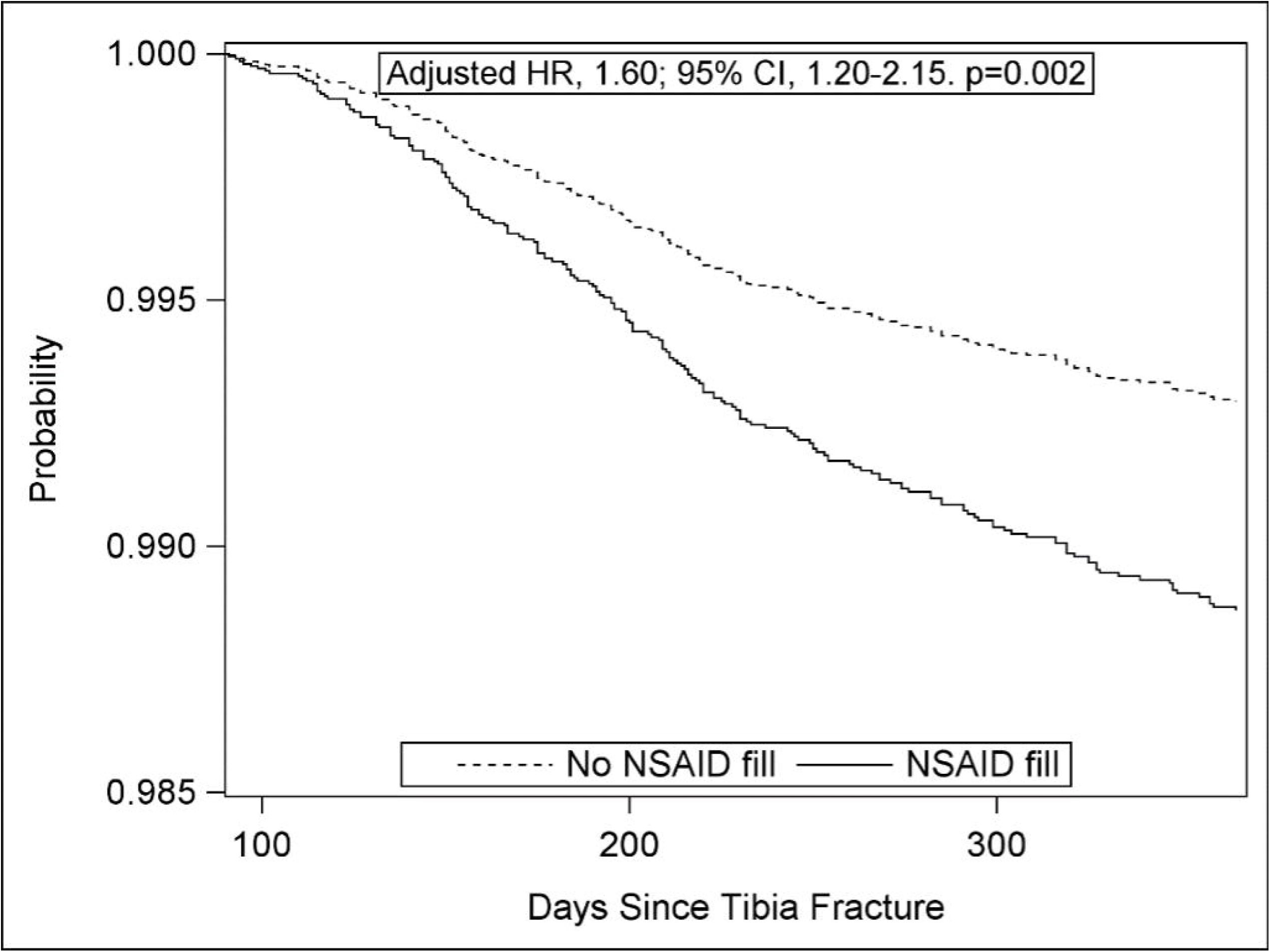

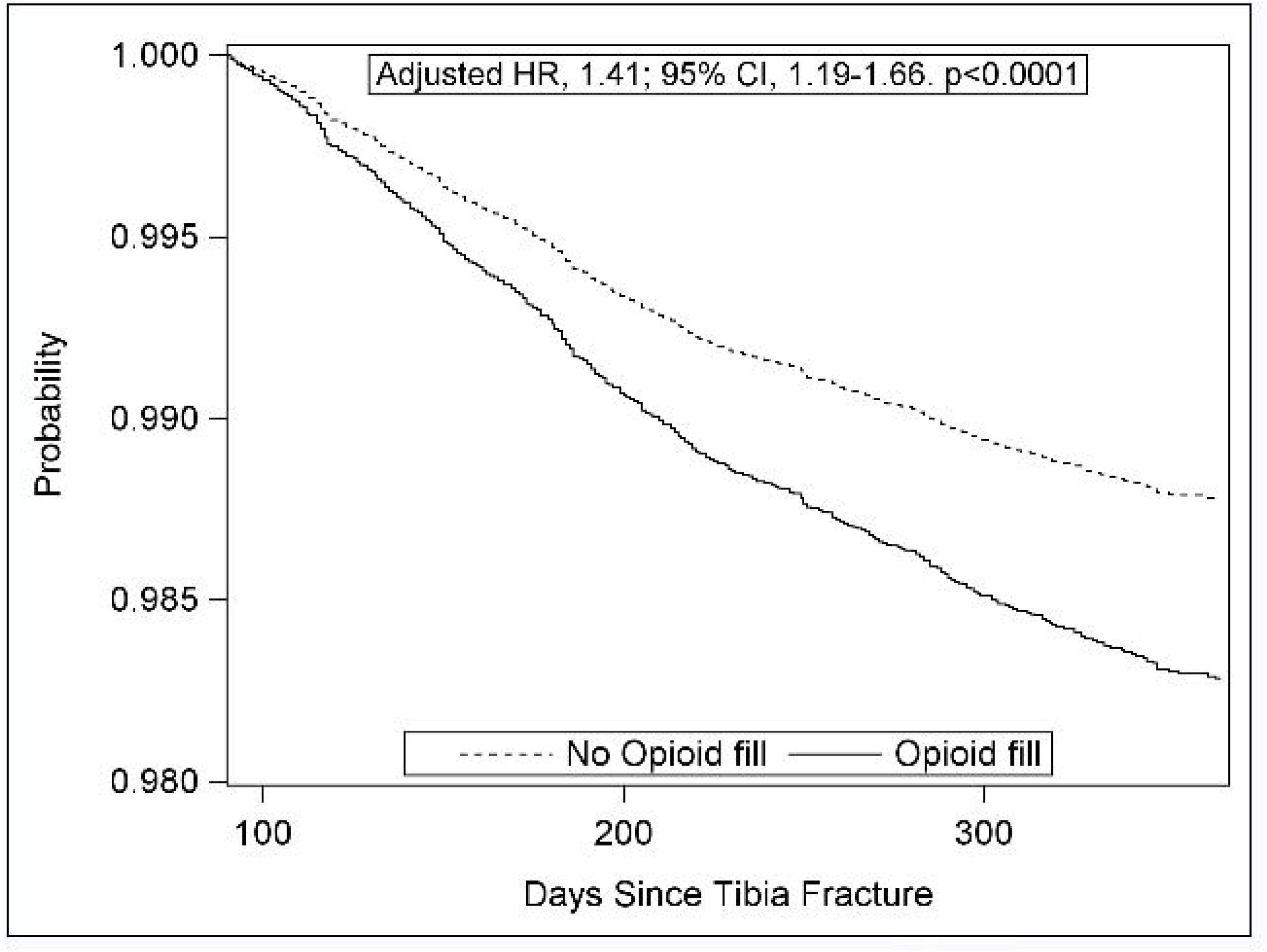

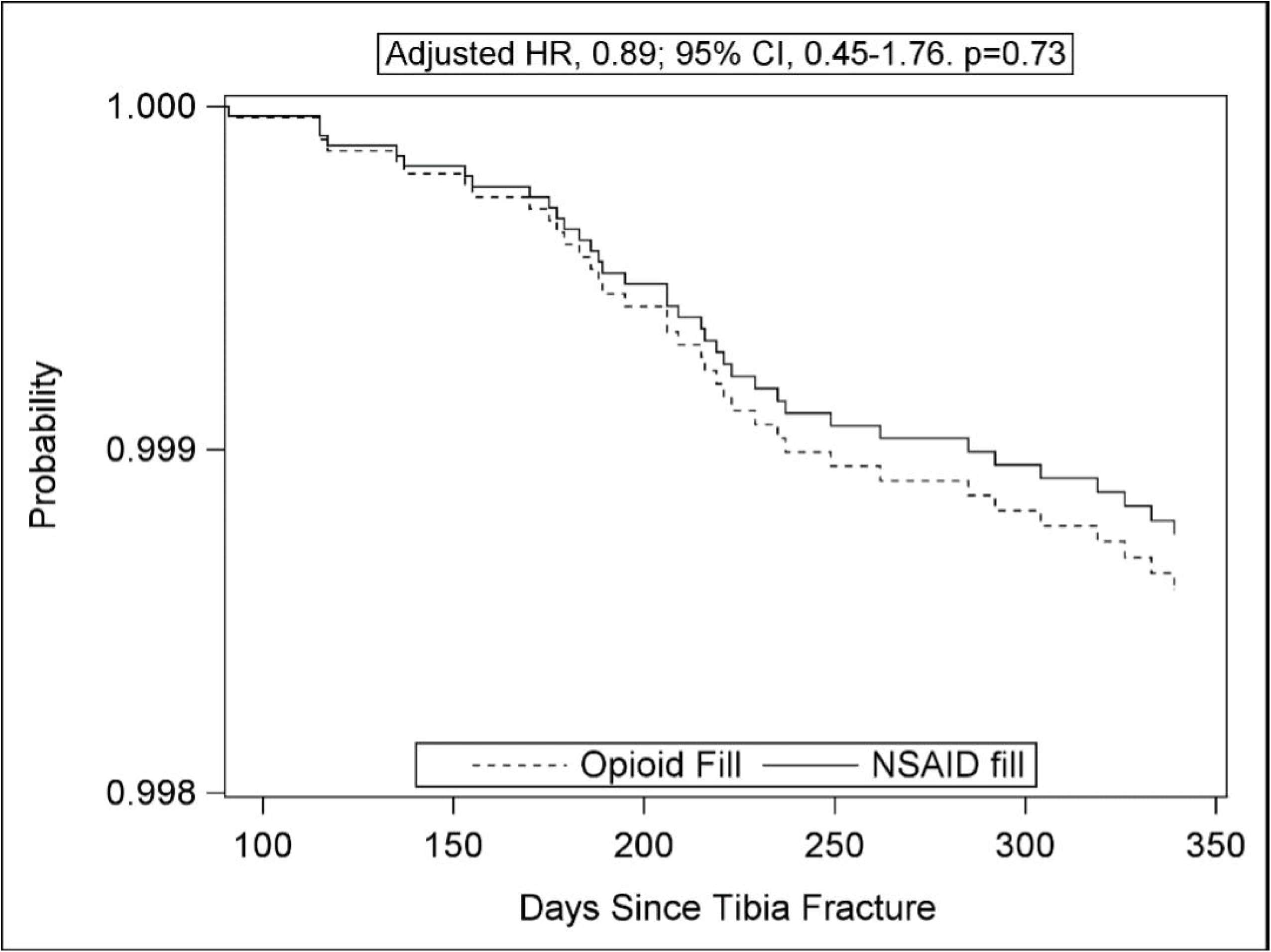

