## Supplemental Table 2 for "Do non-steroidal anti-inflammatory medications or opioids increase the risk of nonunion after surgical and nonsurgical treatment of tibial shaft fractures?"

Supplemental Table 2. Baseline characteristics comparing those who filled an **opioid within 30 days** of tibial fracture versus **those who did not**.

| **Variable (# (%)) unless otherwise specified)** | **Matched Cohort** | |
| --- | --- | --- |
|  | **No Opioid (n=27,046)** | **Opioid (n=27,046)** |
| **Demographics** |  |  |
| Age in years (median (IQR)) | 47 (32, 56) | 47 (33, 56) |
| Male | 13,438 (49.7%) | 13,582 (50.2%) |
| Female | 13,608 (50.3%) | 13,464 (49.8%) |
| Geographic region |  |  |
| North Central | 4,269 (15.8%) | 4,071 (15.1%) |
| South | 6,801 (25.2%) | 6,735 (24.9%) |
| West | 11,063 (40.9%) | 11,487 (42.5%) |
| Northeast | 4,913 (18.2%) | 4,753 (17.8%) |
| Year of fracture exposure (median (IQR)) | 2012 (2010, 2015) | 2012 (2010, 2016) |
| **Exact Matching Variables** |  |  |
| History of tobacco use | 1,425 (5.3%) | 1,425 (5.3%) |
| Open fracture | 3,567 (13.2%) | 3,567 (13.2%) |
| Surgical site infection within 30 days | 110 (0.4%) | 110 (0.4%) |
| ORIF from 0-7 days | 2,485 (9.2%) | 2,485 (9.2%) |
| ORIF from 8-30 days | 271 (1.0%) | 271 (1.0%) |
| **Diagnoses indicating contraindication to NSAID use** **within year prior to fracture** |  |  |
| Chronic kidney disease | 427 (1.6%) | 399 (1.5%) |
| End stage renal disease | 120 (0.4%) | 102 (0.4%) |
| Acute kidney injury (+/- 7 days of fracture) | 324 (1.2%) | 274 (1.0%) |
| Diabetes | 2,832 (10.5%) | 2,889 (10.7%) |
| Cirrhosis | 672 (2.5%) | 662 (2.5%) |
| Peptic ulcer disease | 116 (0.4%) | 117 (0.4%) |
| GI bleed | 159 (0.6%) | 146 (0.5%) |
| Congestive heart failure | 528 (2.0%) | 488 (1.8%) |
| Active anticoagulation | 883 (3.3%) | 833 (3.1%) |
| **Diagnoses indicating increased risk of nonunion** |  |  |
| Chronic steroid use | 379 (1.4%) | 402 (1.5%) |
| Obesity | 1,937 (7.2%) | 1,948 (7.2%) |
| Alcohol use disorder | 69 (0.3%) | 77 (0.3%) |
| History of tibial fracture nonunion | 168 (0.6%) | 149 (0.6%) |
| **Fracture/treatment characteristics** |  |  |
| Internal fixation | 2,752 (10.2%) | 2,741 (10.1%) |
| External fixation | 551 (2.0%) | 756 (2.8%) |
| Casting/nonoperative treatment | 915 (3.4%) | 952 (3.5%) |
| Mechanism of injury: High | 3,090 (11.4%) | 4,047 (15.0%) |
| Mechanism of injury: Low | 1,320 (4.9%) | 1,852 (6.9%) |
| Mechanism of injury: Other | 22,874 (84.6%) | 21,513 (79.5%) |
| **Other diagnoses within year prior to fracture** |  |  |
| Charlson co-morbidity score (median (IQR)) | 0 (0, 1) | 0 (0, 1) |
| 0 | 20,050 (74.1%) | 19,973 (73.9%) |
| 1-2 | 5,372 (19.9%) | 5,556 (20.5%) |
| 3-4 | 1096 (4.1%) | 1010 (3.7%) |
| 5+ | 528 (2.0%) | 507 (1.9%) |
| Generalized anxiety disorder | 734 (2.7%) | 771 (2.9%) |
| Mood disorder | 1,917 (7.1%) | 1,984 (7.3%) |
| Chronic pain | 505 (1.9%) | 495 (1.8%) |
| Coronary artery disease | 912 (3.4%) | 950 (3.5%) |
| Hypertension | 6,625 (24.5%) | 6,868 (25.4%) |
| Cerebrovascular disease | 655 (2.4%) | 615 (2.3%) |
| Inflammatory arthritis | 470 (1.7%) | 487 (1.8%) |
| Peripheral vascular disease | 544 (2.0%) | 526 (1.9%) |
| # of clusters of pain (median (IQR)) | 2 (1, 3) | 1 (0, 3) |
| Opioid use disorder | 1,485 (5.5%) | 1,515 (5.6%) |
| **Pain medication fills in year prior to fracture** |  |  |
| Opioid fill | 8,665 (32.0%) | 8,862 (32.8%) |
| # opioid fills (among those with ≥1) (median (IQR)) | 1 (1, 3) | 1 (1, 3) |
| Morphine milligram equivalents (among those with ≥1 fill) (median (IQR)) | 40 (29, 60) | 38 (27, 58) |
| Duration of opioid (sum of days’ supplies among those with ≥1) (median (IQR)) | 7 (4, 15) | 7 (4, 16) |
| NSAID fill | 6,493 (24.0%) | 6,347 (23.5%) |
| # of NSAID fills (among those with ≥1) (median (IQR)) | 1 (1, 2) | 1 (1, 3) |
| Muscle relaxant fill | 2,541 (9.4%) | 2,731 (10.1%) |
| Acetaminophen fill | 372 (1.4%) | 398 (1.5%) |
| Gabapentin fill | 967 (3.6%) | 1046 (3.9%) |
| Duloxetine fill | 571 (2.1%) | 629 (2.3%) |
| **Pain medication fills in 30 days after fracture** |  |  |
| Opioid fill | 0 (0%) | 27,046 (100%) |
| Muscle relaxant fill | 799 (3.0%) | 1,383 (5.1%) |
| Acetaminophen fill | 106 (0.4%) | 146 (0.5%) |
| Gabapentin fill | 530 (2.0%) | 848 (3.1%) |
| Duloxetine fill | 216 (0.8%) | 269 (1.0%) |

Abbreviations

IQR: Interquartile range

MME: Morphine milligram equivalents

NSAID: Non-steroidal anti-inflammatory drugs

ORIF: Open reduction and internal fixation
