## Supplemental Table 3 for "Do non-steroidal anti-inflammatory medications or opioids increase the risk of nonunion after surgical and nonsurgical treatment of tibial shaft fractures?"

Supplemental Table 3. Outcomes and reasons for end of follow-up in the comparison of those who **filled opioids** within 30 days of tibia fracture versus **those who did not**.

|  | **No Opioid n=27,046** | **Opioid n=27,046** |
| --- | --- | --- |
| Procedure to treat non-union (primary outcome) | 244 (0.9%) | 350 (1.3%) |
| Days to procedure to treat non-union (median, IQR) | 183 (134, 250) | 186 (139, 251) |
| **Reason for end of follow-up** |  |  |
| Enrollment ended before 1 year | 7,121 (26.3%) | 6,780 (25.1%) |
| Reached 1 year without outcome | 19,598 (72.5%) | 19,860 (73.4%) |
| Study ended (Dec 31, 2022) | 83 (0.3%) | 56 (0.2%) |
| Procedure to treat non-union | 244 (0.9%) | 350 (1.3%) |
