## Supplemental Table 4 for "Do non-steroidal anti-inflammatory medications or opioids increase the risk of nonunion after surgical and nonsurgical treatment of tibial shaft fractures?"

Supplemental Table 4. Baseline characteristics comparing those who filled an **NSAID within 30 days** of tibia fracture versus those who filled **opioids within 30 days**.

| **Variable (# (%)) unless otherwise specified)** | **Matched Cohort** | |
| --- | --- | --- |
|  | **Opioid (n=2666)** | **NSAID (n=2666)** |
| **Demographics** |  |  |
| Age in years (median (IQR)) | 49 (36, 57) | 49 (37, 57) |
| Male | 1,170 (43.9%) | 1,157 (43.4%) |
| Female | 1,496 (56.1%) | 1,509 (56.6%) |
| Geographic region |  |  |
| North Central | 478 (17.9%) | 465 (17.4%) |
| South | 511 (19.2%) | 569 (21.3%) |
| West | 1,254 (47.0%) | 1,209 (45.3%) |
| Northeast | 423 (15.9%) | 423 (15.9%) |
| Year of fracture exposure (median (IQR)) | 2014 (2011, 2017) | 2013 (2010, 2017) |
| **Exact Matching Variables** |  |  |
| History of tobacco use | 130 (4.9%) | 130 (4.9%) |
| Open fracture | 183 (6.9%) | 183 (6.9%) |
| Surgical site infection within 90 days | 5 (0.2%) | 5 (0.2%) |
| ORIF from 0-7 days | 123 (4.6%) | 123 (4.6%) |
| ORIF from 8-30 days | 3 (0.1%) | 3 (0.1%) |
| **Diagnoses indicating contraindication to NSAID use** **within year prior to fracture** |  |  |
| Chronic kidney disease | 18 (0.7%) | 21 (0.8%) |
| End stage renal disease | 2 (0.1%) | 2 (0.1%) |
| Acute kidney injury (+/- 7 days of fracture) | 18 (0.7%) | 20 (0.8%) |
| Diabetes | 268 (10.1%) | 291 (10.9%) |
| Cirrhosis | 59 (2.2%) | 65 (2.4%) |
| Peptic ulcer disease | 8 (0.3%) | 5 (0.2%) |
| GI bleed | 10 (0.4%) | 10 (0.4%) |
| Congestive heart failure | 44 (1.7%) | 36 (1.4%) |
| Active anticoagulation | 76 (2.9%) | 89 (3.3%) |
| **Diagnoses indicating increased risk of nonunion** |  |  |
| Chronic steroid use | 43 (1.6%) | 52 (2.0%) |
| Obesity | 258 (9.7%) | 263 (9.9%) |
| Alcohol use disorder | 3 (0.1%) | 3 (0.1%) |
| History of tibial fracture nonunion | 24 (0.9%) | 23 (0.9%) |
| **Fracture/treatment characteristics** |  |  |
| Internal fixation | 126 (4.7%) | 126 (4.7%) |
| External fixation | 11 (0.4%) | 10 (0.4%) |
| Casting/nonoperative treatment | 74 (2.8%) | 85 (3.2%) |
| Mechanism of injury: High | 202 (7.6%) | 203 (7.6%) |
| Mechanism of injury: Low | 122 (4.6%) | 122 (4.6%) |
| Mechanism of injury: Other | 2,361 (88.6%) | 2,363 (88.6%) |
| **Other diagnoses within year prior to fracture** |  |  |
| Charlson co-morbidity score (median (IQR)) | 0 (0, 1) | 0 (0, 1) |
| 0 | 1,914 (71.8%) | 1,889 (70.9%) |
| 1-2 | 610 (22.9%) | 627 (23.5%) |
| 3-4 | 117 (4.4%) | 117 (4.4%) |
| 5+ | 25 (0.9%) | 33 (1.2%) |
| Generalized anxiety disorder | 92 (3.5%) | 94 (3.5%) |
| Mood disorder | 214 (8.0%) | 227 (8.5%) |
| Chronic pain | 91 (3.4%) | 90 (3.4%) |
| Coronary artery disease | 92 (3.5%) | 80 (3.0%) |
| Hypertension | 771 (28.9%) | 798 (29.9%) |
| Cerebrovascular disease | 62 (2.3%) | 61 (2.3%) |
| Inflammatory arthritis | 98 (3.7%) | 97 (3.6%) |
| Peripheral vascular disease | 47 (1.8%) | 49 (1.8%) |
| # of clusters of pain (median (IQR)) | 2 (1, 3) | 2 (1, 3) |
| Opioid use disorder | 179 (6.7%) | 194 (7.3%) |
| **Pain medication fills in year prior to fracture** |  |  |
| Opioid fill | 1,036 (38.9%) | 1,048 (39.3%) |
| # opioid fills (among those with ≥1) (median (IQR)) | 2 (1, 3) | 1 (1, 3) |
| Morphine milligram equivalents (among those with ≥1 fill) (median (IQR)) | 36 (25, 52) | 37 (26, 55) |
| Duration of opioid (sum of days’ supplies among those with ≥1) (median (IQR)) | 6 (3, 15) | 7 (4, 18) |
| NSAID fill | 1,156 (43.4%) | 1,374 (51.5%) |
| # of NSAID fills (among those with ≥1) (median (IQR)) | 3 (1, 6) | 2 (1, 5) |
| Muscle relaxant fill | 376 (14.1%) | 383 (14.4%) |
| Acetaminophen fill | 59 (2.2%) | 62 (2.3%) |
| Gabapentin fill | 166 (6.2%) | 166 (6.2%) |
| Duloxetine fill | 66 (2.5%) | 81 (3.0%) |
| **Pain medication fills in 30 days after fracture** |  |  |
| Opioid fill | 2,666 (100%) | 0 (0.0%) |
| Muscle relaxant fill | 224 (8.4%) | 220 (8.3%) |
| Acetaminophen fill | 30 (1.1%) | 30 (1.1%) |
| Gabapentin fill | 128 (4.8%) | 117 (4.4%) |
| Duloxetine fill | 43 (1.6%) | 51 (1.9%) |

Abbreviations

IQR: Interquartile range

MME: Morphine milligram equivalents

NSAID: Non-steroidal anti-inflammatory drugs

ORIF: Open reduction and internal fixation
