## Supplemental Table 5 for "Do non-steroidal anti-inflammatory medications or opioids increase the risk of nonunion after surgical and nonsurgical treatment of tibial shaft fractures?"

Supplemental Table 5. Outcomes and reasons for end of follow-up in the comparison of those who **filled NSAIDs** versus those who **filled opioids** within 30 days of tibia fracture.

|  | **Opioid n=2666** | **NSAID n=2666** |
| --- | --- | --- |
| Procedure to treat non-union (primary outcome) | 18 (0.7%) | 19 (0.7%) |
| Days to procedure to treat non-union (median, IQR) | 220 (153, 292) | 195 (175, 237) |
| **Reason for end of follow-up** |  |  |
| Enrollment ended before 1 year | 723 (27.1%) | 660 (24.8%) |
| Reached 1 year without outcome | 1,916 (71.9%) | 1,969 (73.9%) |
| Study ended (Dec 31, 2022) | 9 (0.3%) | 18 (0.7%) |
| Non-union treatment | 18 (0.7%) | 19 (0.7%) |
