## Supplemental Table 6 for "Do non-steroidal anti-inflammatory medications or opioids increase the risk of nonunion after surgical and nonsurgical treatment of tibial shaft fractures?"

Supplemental Table 6. Baseline characteristics comparing of those who **filled ≥2 NSAIDs** within 30 days versus **those who did not fill any NSAIDs**.

| **Variable (# (%)) unless otherwise specified)** | **Matched Cohort** | |
| --- | --- | --- |
|  | **No NSAID (n=996)** | **≥2 NSAIDs (n=996)** |
| **Demographics** |  |  |
| Age in years (median (IQR)) | 45 (32, 55) | 45 (33, 55) |
| Male | 484 (48.6%) | 489 (49.1%) |
| Female | 512 (51.4%) | 507 (50.9%) |
| Geographic region |  |  |
| North Central | 128 (12.9%) | 133 (13.4%) |
| South | 197 (19.8%) | 197 (19.8%) |
| West | 528 (53.0%) | 509 (51.1%) |
| Northeast | 143 (14.4%) | 157 (15.8%) |
| Year of fracture exposure (median (IQR)) | 2014 (2011, 2019) | 2014 (2011, 2019) |
| **Exact Matching Variables** | 60 (6.0%) | 60 (6.0%) |
| History of tobacco use | 98 (9.8%) | 98 (9.8%) |
| Open fracture | 4 (0.4%) | 4 (0.4%) |
| Surgical site infection within 30 days |  |  |
| MME from 0-30 days |  |  |
| 0 | 182 (18.3%) | 182 (18.3%) |
| >0 to <45 | 372 (37.3%) | 372 (37.3%) |
| ≥45 to <75 | 239 (24.0%) | 239 (24.0%) |
| ≥75 | 203 (20.4%) | 203 (20.4%) |
| # of opioid fills from 0-30 days |  |  |
| 0 | 181 (18.2%) | 181 (18.2%) |
| 1 | 243 (24.4%) | 243 (24.4%) |
| >1 | 572 (57.4%) | 572 (57.4%) |
| ORIF from 0-7 days | 159 (16.0%) | 159 (16.0%) |
| ORIF from 8-30 days | 27 (2.7%) | 27 (2.7%) |
| **Diagnoses indicating contraindication to NSAID use** **within year prior to fracture** |  |  |
| Chronic kidney disease | 12 (1.2%) | 7 (0.7%) |
| End stage renal disease | 3 (0.3%) | 1 (0.1%) |
| Acute kidney injury (+/- 7 days of fracture) | 8 (0.8%) | 8 (0.8%) |
| Diabetes | 60 (6.0%) | 77 (7.7%) |
| Cirrhosis | 19 (1.9%) | 22 (2.2%) |
| Peptic ulcer disease | 1 (0.1%) | 1 (0.1%) |
| GI bleed | 7 (0.7%) | 6 (0.6%) |
| Congestive heart failure | 4 (0.4%) | 6 (0.6%) |
| Active anticoagulation | 14 (1.4%) | 15 (1.5%) |
| **Diagnoses indicating increased risk of nonunion** |  |  |
| Chronic steroid use | 14 (1.4%) | 15 (1.5%) |
| Obesity | 96 (9.6%) | 97 (9.7%) |
| Alcohol use disorder | 1 (0.1%) | 2 (0.2%) |
| History of tibial fracture nonunion | 0 (0.0%) | 1 (0.1%) |
| **Fracture/treatment characteristics** |  |  |
| Internal fixation | 188 (18.9%) | 188 (18.9%) |
| External fixation | 13 (1.3%) | 16 (1.6%) |
| Casting/nonoperative treatment | 33 (3.3%) | 36 (3.6%) |
| Mechanism of injury: High | 190 (19.1%) | 189 (19.0%) |
| Mechanism of injury: Low | 149 (15.0%) | 145 (14.6%) |
| Mechanism of injury: Other | 686 (68.9%) | 691 (69.4%) |
| **Other diagnoses within year prior to fracture** |  |  |
| Charlson co-morbidity score (median (IQR)) | 0 (0, 0) | 0 (0, 0) |
| 0 | 792 (79.5%) | 764 (76.7%) |
| 1-2 | 162 (16.3%) | 195 (19.6%) |
| 3-4 | 31 (3.1%) | 26 (2.6%) |
| 5+ | 11 (1.1%) | 11 (1.1%) |
| Generalized anxiety disorder | 29 (2.9%) | 36 (3.6%) |
| Mood disorder | 79 (7.9%) | 87 (8.7%) |
| Chronic pain | 24 (2.4%) | 31 (3.1%) |
| Coronary artery disease | 14 (1.4%) | 19 (1.9%) |
| Hypertension | 218 (21.9%) | 248 (24.9%) |
| Cerebrovascular disease | 18 (1.8%) | 14 (1.4%) |
| Inflammatory arthritis | 19 (1.9%) | 18 (1.8%) |
| Peripheral vascular disease | 12 (1.2%) | 16 (1.6%) |
| # of clusters of pain (median (IQR)) | 1 (0, 3) | 1 (0, 3) |
| Opioid use disorder | 56 (5.6%) | 70 (7.0%) |
| **Pain medication fills in year prior to fracture** |  |  |
| Opioid fill | 327 (32.8%) | 342 (34.3%) |
| # opioid fills (among those with ≥1) (median (IQR)) | 2 (1, 4) | 2 (1, 4) |
| Morphine milligram equivalents (among those with ≥1 fill) (median (IQR)) | 37 (23, 53) | 35 (25, 51) |
| Duration of opioid (sum of days’ supplies among those with ≥1) (median (IQR)) | 8 (4, 22) | 7 (3, 21) |
| NSAID fill | 271 (27.2%) | 319 (32.0%) |
| # of NSAID fills (among those with ≥1) (median (IQR)) | 2 (1, 3) | 2 (1, 3) |
| Muscle relaxant fill | 137 (13.8%) | 139 (14.0%) |
| Acetaminophen fill | 25 (2.5%) | 20 (2.0%) |
| Gabapentin fill | 49 (4.9%) | 52 (5.2%) |
| Duloxetine fill | 18 (1.8%) | 29 (2.9%) |
| **Pain medication fills in 30 days after fracture** |  |  |
| Opioid fill | 815 (81.8%) | 815 (81.8%) |
| Muscle relaxant fill | 180 (18.1%) | 164 (16.5%) |
| Acetaminophen fill | 22 (2.2%) | 16 (1.6%) |
| Gabapentin fill | 128 (12.9%) | 123 (12.3%) |
| Duloxetine fill | 15 (1.5%) | 16 (1.6%) |

Abbreviations

IQR: Interquartile range

MME: Morphine milligram equivalents

NSAID: Non-steroidal anti-inflammatory drugs

ORIF: Open reduction and internal fixation
