## Supplementary figures and images for "Do non-steroidal anti-inflammatory medications or opioids increase the risk of nonunion after surgical and nonsurgical treatment of tibial shaft fractures?"

### Supplemental Figure 1

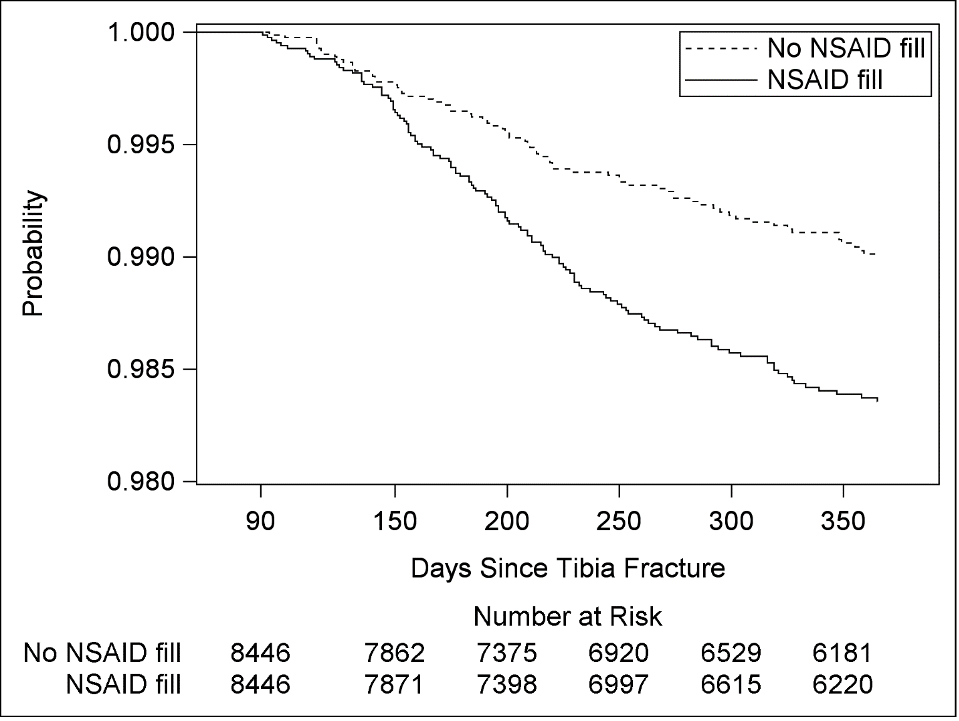

### Supplemental Figure 2

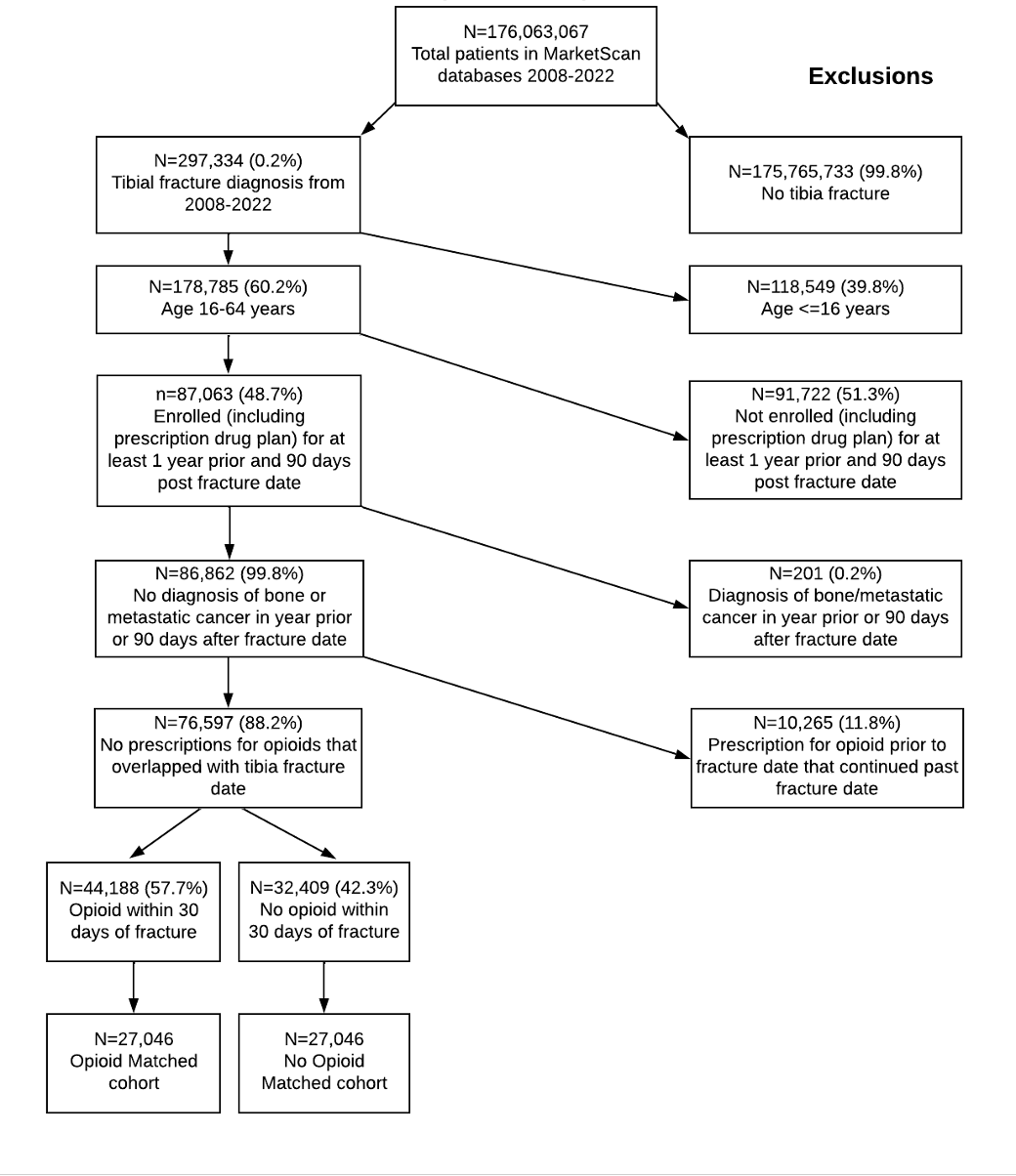

### Supplemental Figure 3

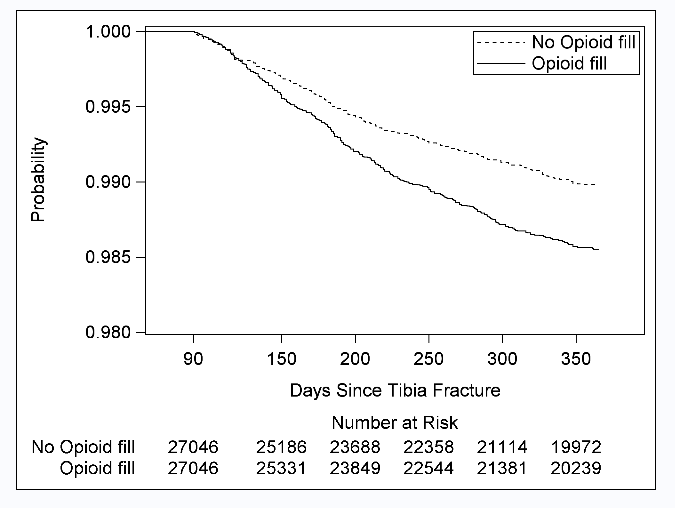

### Supplemental Figure 4

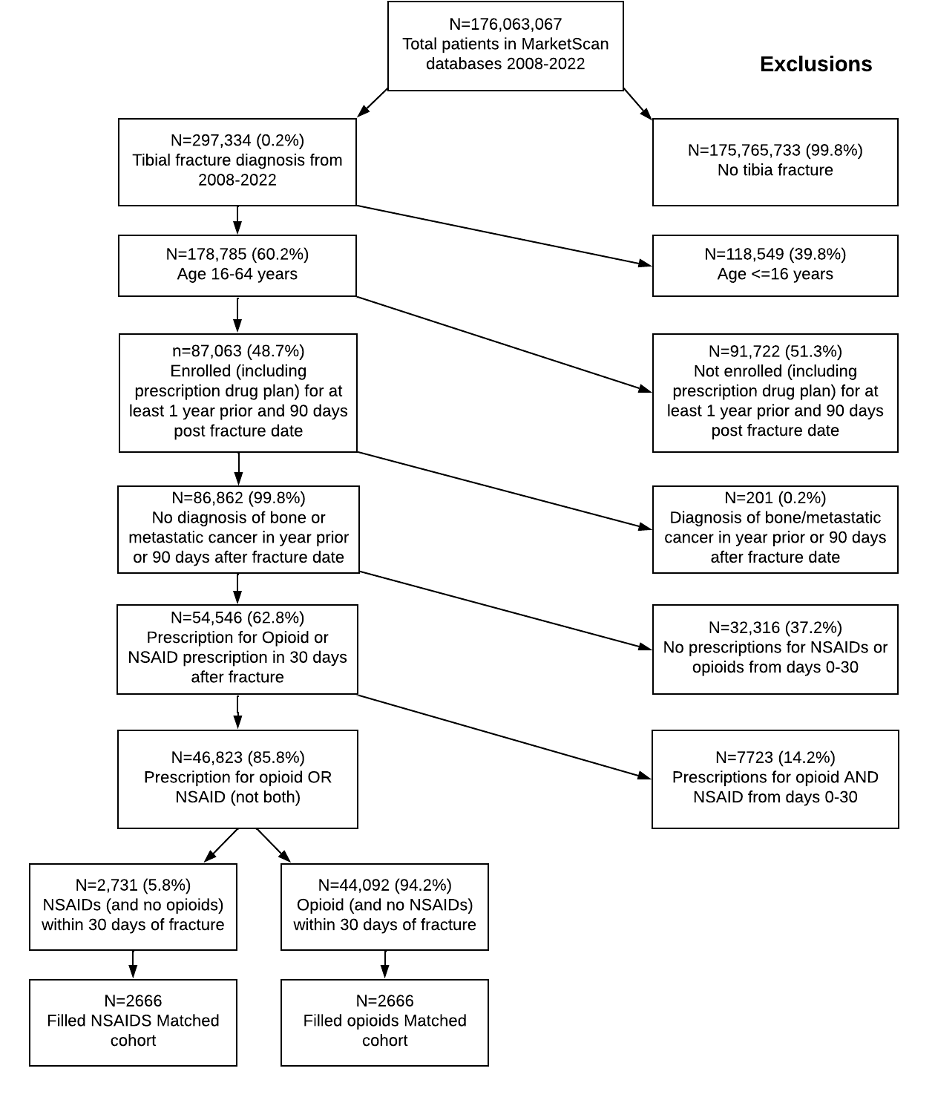

### Supplemental Figure 5

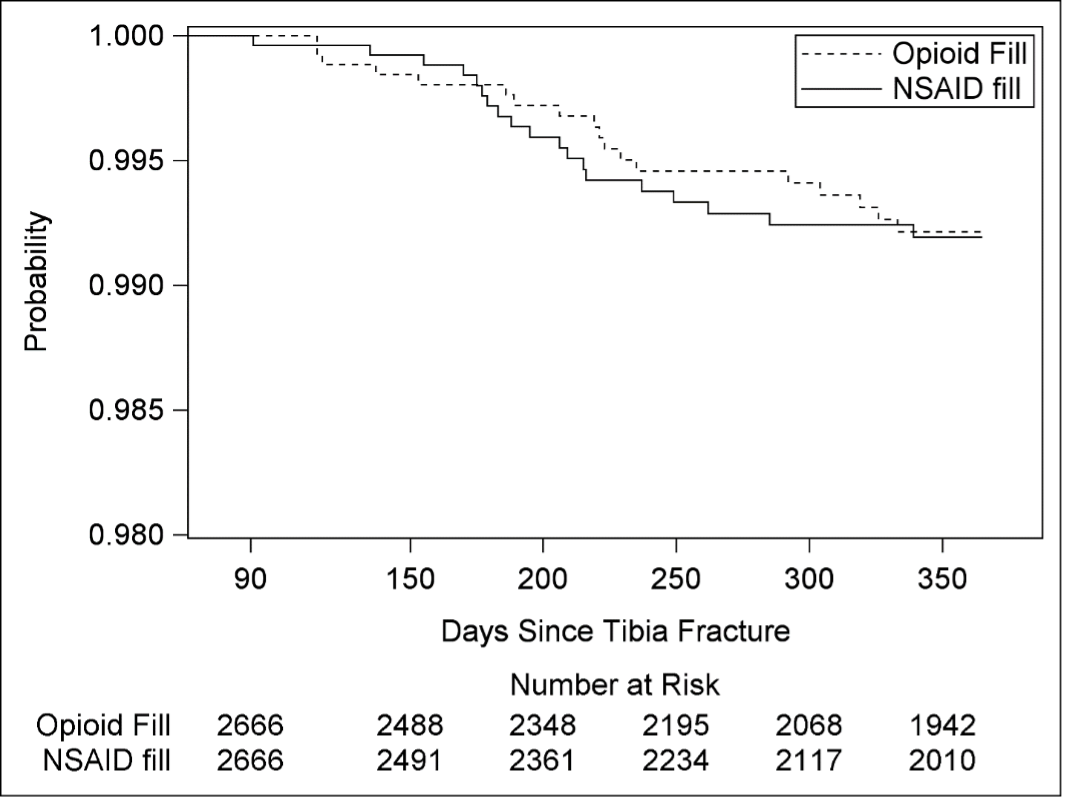

### Supplemental Figure 6

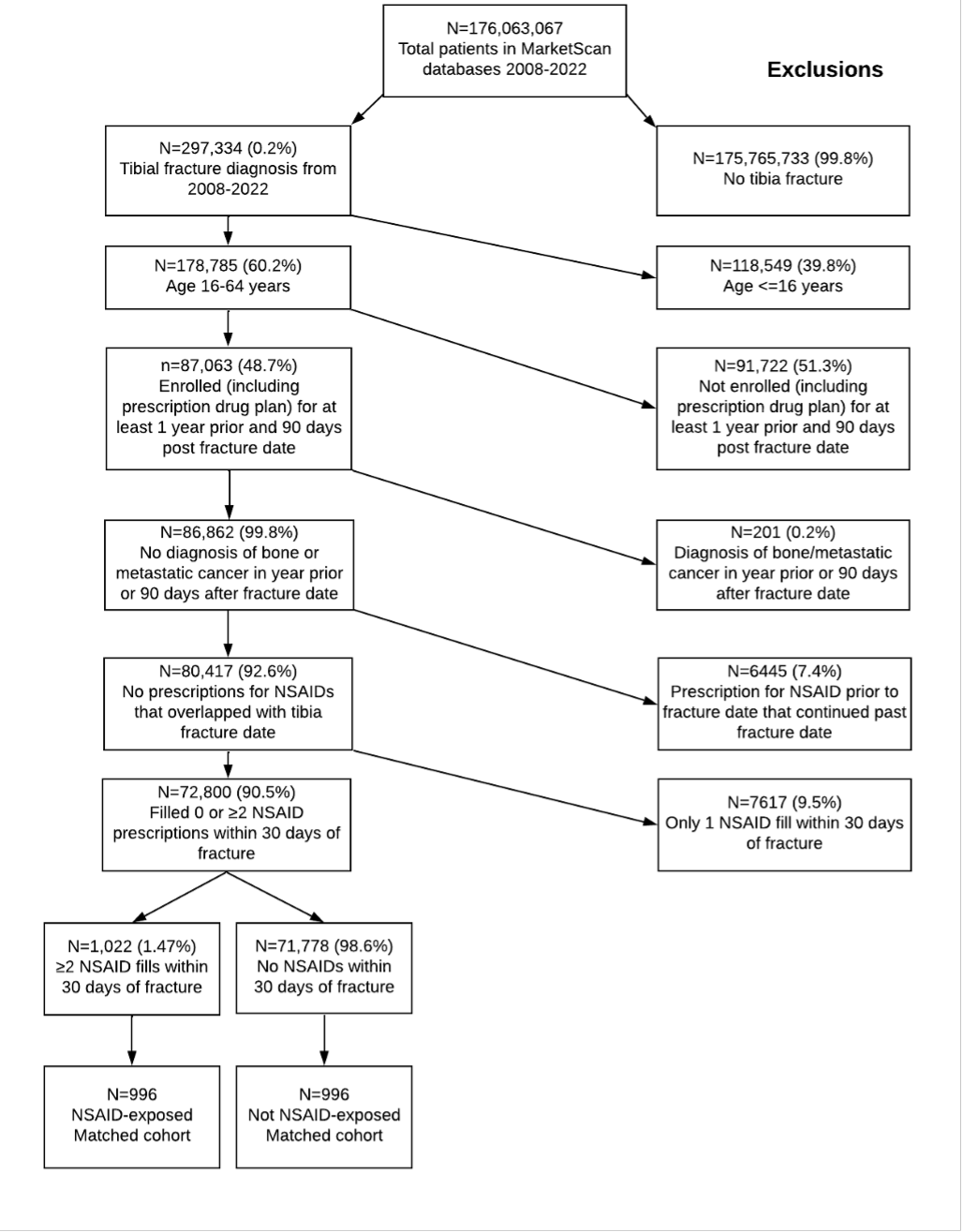

### Supplemental Figure 7

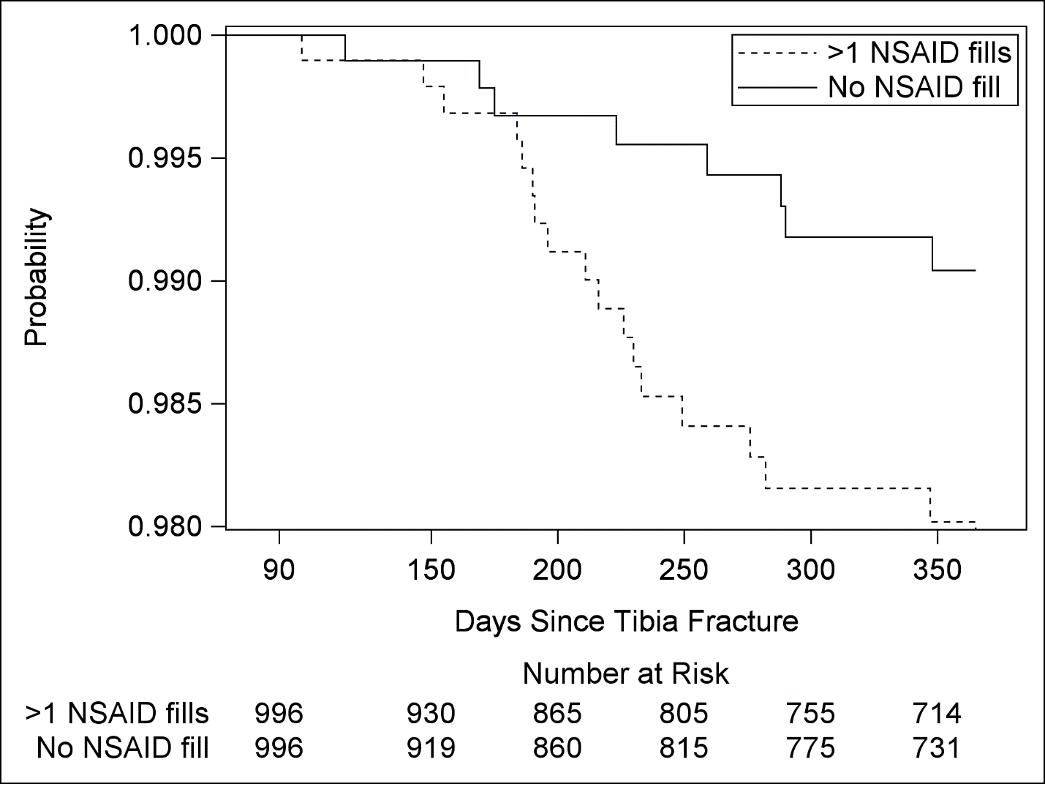

### Supplemental Figure 8

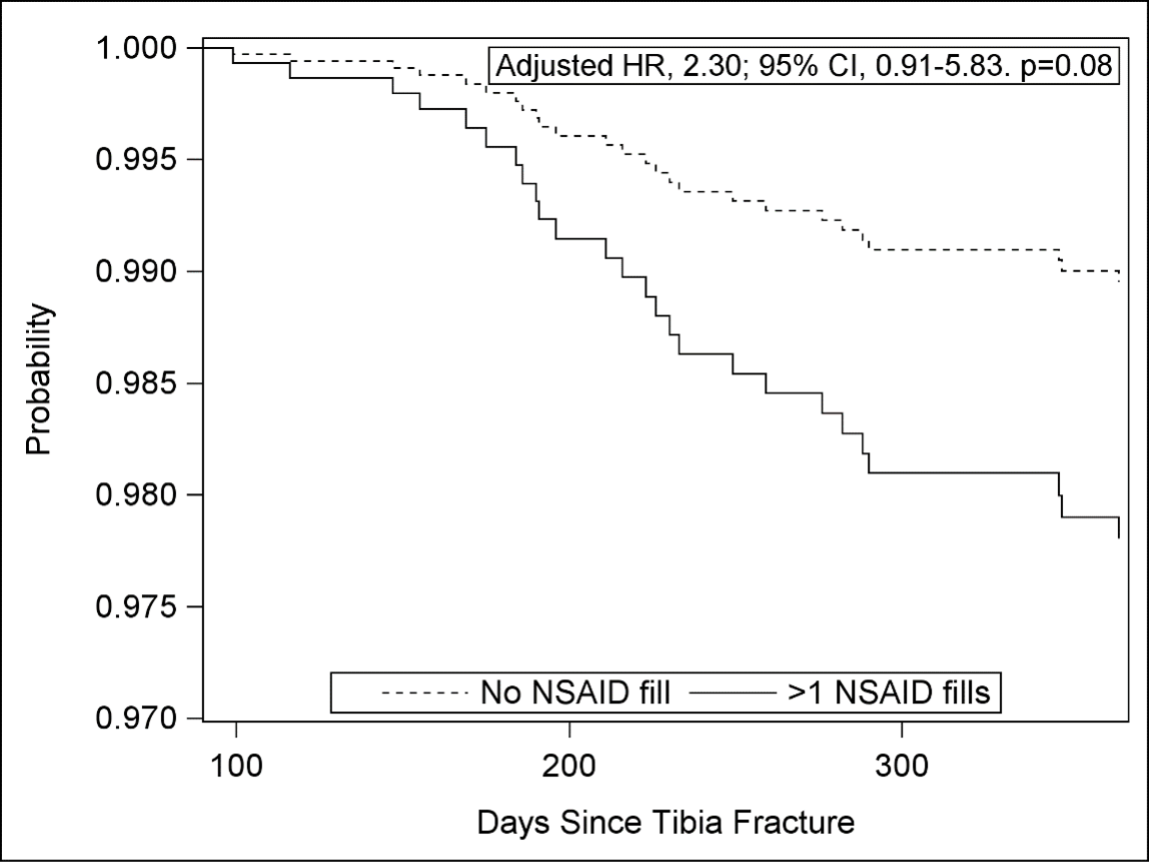
