## Supplemental Table 7 for "Do non-steroidal anti-inflammatory medications or opioids increase the risk of nonunion after surgical and nonsurgical treatment of tibial shaft fractures?"

Supplemental Table 7. Outcomes and reasons for end of follow-up in the comparison of those who **filled ≥2 NSAIDs** within 30 days versus **those who did not fill any NSAIDs**.

|  | **No NSAID n=996** | **NSAID n=996** |
| --- | --- | --- |
| Procedure to treat non-union (primary outcome) | 8 (0.8%) | 18 (1.8%) |
| Days to procedure to treat non-union (median, IQR) | 241 (172, 289) | 213 (186, 249) |
| **Reason for end of follow-up** |  |  |
| Enrollment ended before 1 year | 267 (26.8%) | 267 (26.8%) |
| Reached 1 year without outcome | 716 (71.9%) | 704 (70.7%) |
| Study ended (Dec 31, 2022) | 5 (0.5%) | 7 (0.7%) |
| Non-union treatment | 8 (0.8%) | 18 (1.8%) |
